# The global effects of alcohol consumption on Gross Domestic Product in high- and low-income countries: a systematic review and meta-analysis

**DOI:** 10.1101/2022.04.27.22274363

**Authors:** Swettha Mahesarajah, Raha Pazoki

## Abstract

**Aims and objectives:** This study aims to measure the disease burden and the economic burden associated with alcohol consumption in both high- and low-income countries. To emphasise the necessity of making this issue a worldwide priority, the percentage of GDP attributable to alcohol-related costs will be stated.

**Design:** Systematic review and meta-analysis

**Data sources:** A systematic search concerning health and social costs was conducted primarily through PubMed and subsequent citation chaining of appropriate systematic reviews. Other electronic databases such as Google Scholar was also freely searched.

**Eligibility criteria for selecting studies:** Observational studies examining alcohol-related harm, alcohol-related disease, and alcohol-related expenditure with all studies measuring alcohol-related harm using the alcohol-attributable fraction (AAF).

**Results:** 9 cross-sectional studies were obtained assessing the consequences of alcohol on the respective country’s economy with all studies utilising a prevalence-based approach. 5 studies were eligible for a meta-analysis in the statistically programming software, R. The pooled estimate of the economic burden of alcohol in 5 countries equated to be 0.01% of GDP. Though inconsistencies in cost estimations resulted in an underestimation, our results provide evidence to suggest that alcohol negatively affects both individuals and society. The available literature on the topic of the economic impact of alcohol is inadequate; especially when investigating concerns in poorer regions of the world.

**Conclusion:** Though the current estimate of global GDP attributable to alcohol use is low, the evidence suggesting the global increase of alcohol consumption is paramount to avoid future calamities. Cooperative leadership from the World Health Organisation (WHO), International Monetary Fund, and the World Bank are requisite to control the harmful patterns of alcohol consumption seen across the globe.

**STRENGTHS AND LIMITATIONS OF THIS STUDY:**

1. This is the first systematic review assessing the detrimental consequences of alcohol on economic health with the inclusion of both lower-middle income and high-income countries.
2. This study provides a pooled estimate of the global estimate of the percentage of GDP attributable to alcohol related costs using statistical package, R which has not been done before.
3. The obtainment of research conducted in low-income countries proved to be difficult, and as a result no low-middle income countries were used when calculating the pooled estimate. Therefore, the accuracy of the provided estimate was decreased.

## INTRODUCTION

Alcohol continues to be one of the main contributors to the global disease burden making its mark as one of the top ten leading risk factors to cause disease and disability (1). The development of mental diseases such as alcohol addiction, anxiety, and depression, are all health implications associated with hazardous alcohol use (2). Another important health implication of alcohol misuse is alcoholic liver disease (ALD) which is becoming more common globally, especially in high-income countries (HICs). The manifestations of ALD caused by excessive drinking include steatosis (fatty liver), alcoholic hepatitis, fibrosis, cirrhosis and lastly, hepatocellular carcinoma (HCC) (3). Furthermore, drinking alcohol increases the risk of over 60 diseases, including myocardial infarction, liver cirrhosis, malignancies (including the liver, larynx, oesophagus, and colon), haemorrhagic stroke, chronic respiratory disorders, and diabetes (4, 5). These being possible causes for 5.3% of all deaths worldwide and 5.1% of all disease/injury with alcohol being a component cause (6). Understanding the significant repercussions of alcohol consumption at the histopathological level and the benefits achieved by the reduction of this lifestyle factor is requisite to aid research and advancements in science with regards to preventable illnesses (7). Nevertheless, estimating the economic burden of alcohol is essential to make accurate decisions about resource allocation and effectual policy making.

Gross domestic product (GDP) is a measure of a country’s economic health, it is the sum of final consumption, investment, and net exports. It stands as a good indicator for the economic size and overall performance of a country (8). To better understand prices on healthcare spending across developing and developed countries as well as how the reduction of alcoholism can alleviate pressures on healthcare expenditure in HICs and LICs, the relationship between alcohol-attributable diseases/injury and the GDP-Purchasing Parity (GDP PPP) could be a possible outlet. Baumberg and colleagues estimated that the global economic burden of alcohol consumption stands at approximately 0.6-2.0% of global GDP. This includes $40-105 billion dollars spent towards alcohol-related health issues, $30-85 billion for crime and violence, and around $0-80 billion towards unemployment (7). The surge in alcohol affordability over the past years poses immense pressures on public health systems and economies (9). A study conducted in the United Kingdom by Lister et al. revealed the economic value per quality-adjusted life year (QALY) for alcohol misuse to be approximately £65,000 for alcohol misuse, whilst cardiovascular diseases held less than half this figure. The main justifications for this are the fact that alcohol misuse brings about elevated exterior costs (10).

Alcohol use disorder (AUD) is the third and fourth most disabling disease category in HICs and low and middle-income countries (LMICs), respectively. AUDs constitute 18.4 million years of life lost to disability (YLDs) in low- and middle-income countries and 3.9 million YLDs in high-income countries (11). Studies have shown that HICs countries have a higher percentage of alcohol consumed per capita than low-income countries (LICs), however, in low socioeconomic statuses, the patterns of alcohol consumption are more dangerous with LICs having substantially higher rates of alcohol-related mortality and morbidity (12). Albeit HICs currently have the biggest alcohol consumption per capita in comparison to LICs and middle-income countries (MICs), alcohol consumption is still gradually increasing in several LMICs (13). The rising prevalence of chronic diseases and the rise in alcohol consumption per capita in developing countries is becoming increasingly alarming (14). Only two studies have been undertaken so far to estimate the economic impact caused by alcohol consumption, and neither study included low-income, lower middle-income, or upper middle-income countries (7,15). This current study aims to systematically review the available literature on the financial implications of alcohol-related disease and injury in high and low-income countries and to provide an estimate for the percentage of GDP attributable to alcohol-related costs to society. This study seeks to provide valuable insights into the negative effects of alcohol on society, as well provide suggestions for public health policy implementation and resource allocation.

## METHODS

### Patient and public involvement

The research process did not involve patient or public involvement.

### Eligibility criteria

The studies presented in the systematic review and later meta-analysis consist of alcohol-related economic costs and state an estimate for the percentage of GDP attributable to alcohol. There was no set age limit defined in the criteria. It was specified that both male and female alcohol-related statistics should be noted for successive calculations. The eligibility criteria (Figure 1) also ensured that the systematic review would consist of only observational study designs, the majority of these being cross-sectional studies. This allowed the prevalence of alcohol-related diseases to be studied at a specific point in time, as well as how alcohol may affect the health of a population (16).

**Figure 1:**
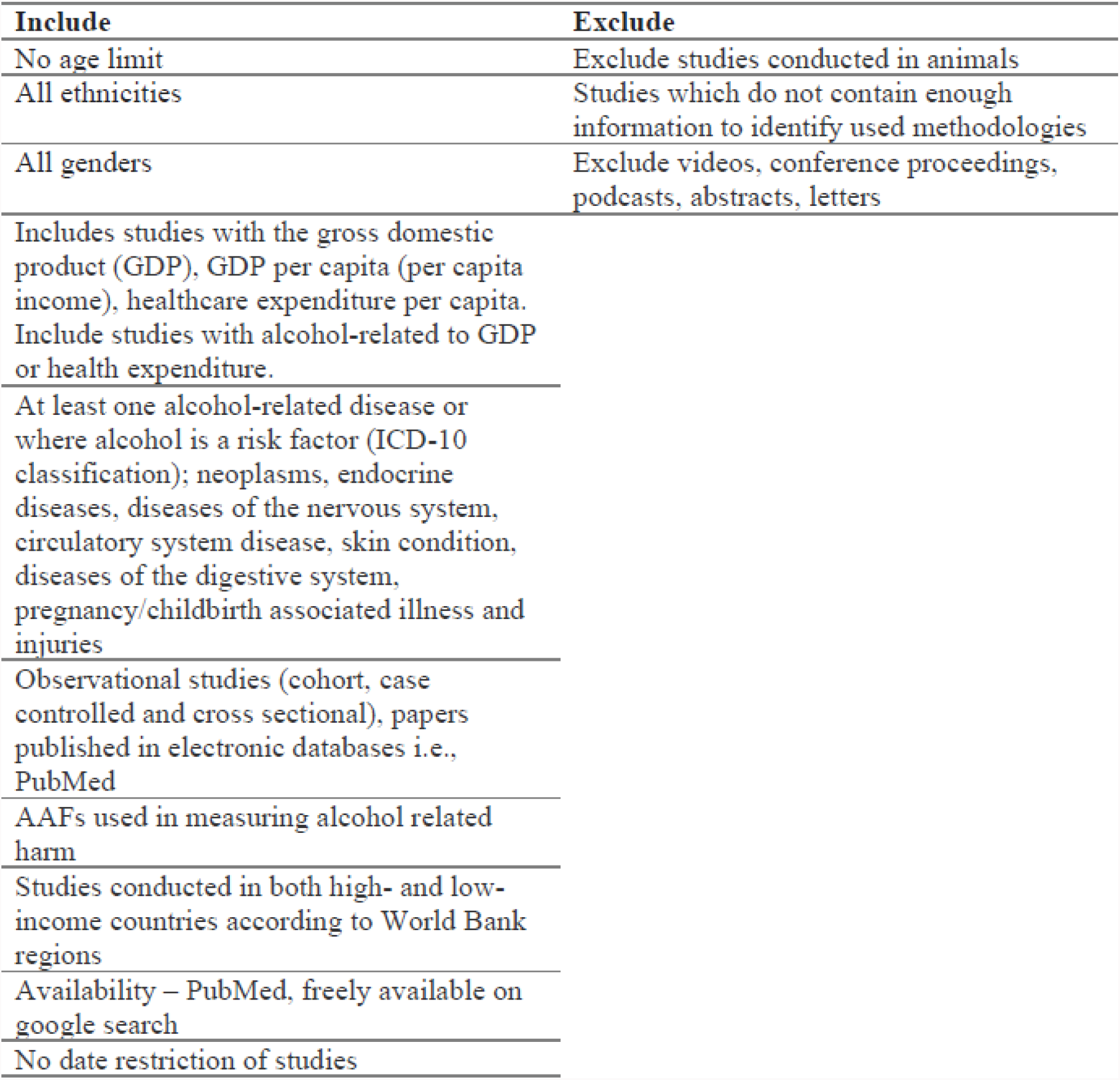
The eligibility criteria. These criteria were created to ensure all studies included maintained uniformity and relevant data. Studies excluded did not meet at least one of the inclusion criteria or met one of the exclusion criteria. All studies selected for the systematic review matched all requirements of the inclusion criteria.

### Literature searching and screening

During the preliminary search of the literature, applicable keywords from articles were identified and noted. This permitted the strategic combination of various terms to provide numerous search strings; these were tested and refined accordingly to produce the final set of search terms. The search terms “alcohol consumption”, “Gross Domestic Product”, “healthcare expenditure”, and “economic cost” were used during the search. Boolean Logic was used to combine and exclude certain key terms, the combination of connectives such as “AND” and “OR” aided the narrowing of results and thus allowing a more productive literature search (17). Medical Subject Headings (MeSH) terms were used when testing search strategies. In the months of 20th January 2021 and 5th February 2021, a systematic search of available literature was performed in PubMed. Google Scholar was also freely searched using search terms combinations including “Gross Domestic Product”, “economic cost”, “alcohol consumption”, “alcohol intake”, and “alcohol harm”, similar systematic reviews were identified, and consequent citation chaining was performed. To avoid missing studies not available through reputable journals, The Journal of Negative Results in Biomedicine (JNRBM 2002-2017) was searched to find relevant unpublished/unpopular articles; a study conducted in Belgium was successfully obtained. The title and abstract were screened per the eligibility criteria, studies which deemed irrelevant were excluded. The full text of the remaining possibly appropriate studies was obtained, again these studies were scrutinised with the aid of the inclusion/exclusion criteria. A total of 9 studies met the entire eligibility criteria.

### Data extraction

A PRISMA flow diagram was utilised to record the number of studies included and excluded at each stage of the searching and screening process (18). Upon submitting the final search string into PubMed, the search results were imported into Mendeley. The number of studies obtained via citation chaining and free searching were also included. The removal of duplicates was completed by both the service provided by Mendeley and manually. The title and abstract of the selected papers were screened on Mendeley most papers were excluded based on their title, however where titles were not clear, the abstract was read to make a decision. Studies that met at least one of the exclusion criteria were eliminated; the paper’s information and reasons for exclusion were stored in a spreadsheet for reproducibility purposes. Full texts of papers that were not available through PubMed were obtained through the google search or journal websites. To reduce bias and enhance validity, a data extraction form was designed to deliver uniformity in the systematic review and meta-analysis. This form enabled the most significant and useful information to be stored in an organised manner for the initial analysis and later meta-analysis. The categories present in this form include bibliographic information, location of study, study design, the alcohol-related diseases specified in the paper, income classification, statistical methods used, study aims and measured outcomes. Total net costs in studies were converted to USD to maintain uniformity and allow for accurate calculations. The selected countries were categorised by income according to the GDP per capita (current USD$) World Bank development Indicator (WDI). To check whether bias was present, sample size and selection of participants was considered, the method of outcome collection and verification was also studied where possible in selected studies to identify the presence of detection bias (19).

### Statistical procedures and data analysis

The random effects model was used in the statistical programming software, R. The statistical procedures allowed pooled estimate computation using the DerSimonian and Liard Method, a variation of inverse-variance method (IVW). Heterogeneity assessment was also conducted using Cochran’s Q index, as well as tests for publication bias. The software packages “robumeta”, “metaphor” and “dplyr” were used when conducting this statistical analysis. A forest plot was generated to display the results of the random effects meta-analysis and a funnel plot was generated to illustrate heterogeneity and detect the possibility for publication bias.

## RESULTS

### Systematic review

The search strategy used for the systematic literature search generated 106 studies in PubMed; a further 20 studies were identified through other sources (freely on google search and bio archives). Of these 126 citations, 61 papers were eliminated following the judgment of their corresponding title and abstract. It was only possible to obtain the full online texts of 41 studies; 1 study was retrieved after subsequent contact with an expert. The majority of papers were excluded based on the fact the study design was not desired on this occasion. These research papers were examined in compliance with the eligibility criteria, 9 studies met the requirements and were therefore used in the analysis. The information concerning the systematic selection of research studies is illustrated in a PRISMA flow chart seen in figure 2.

**Figure 2:**
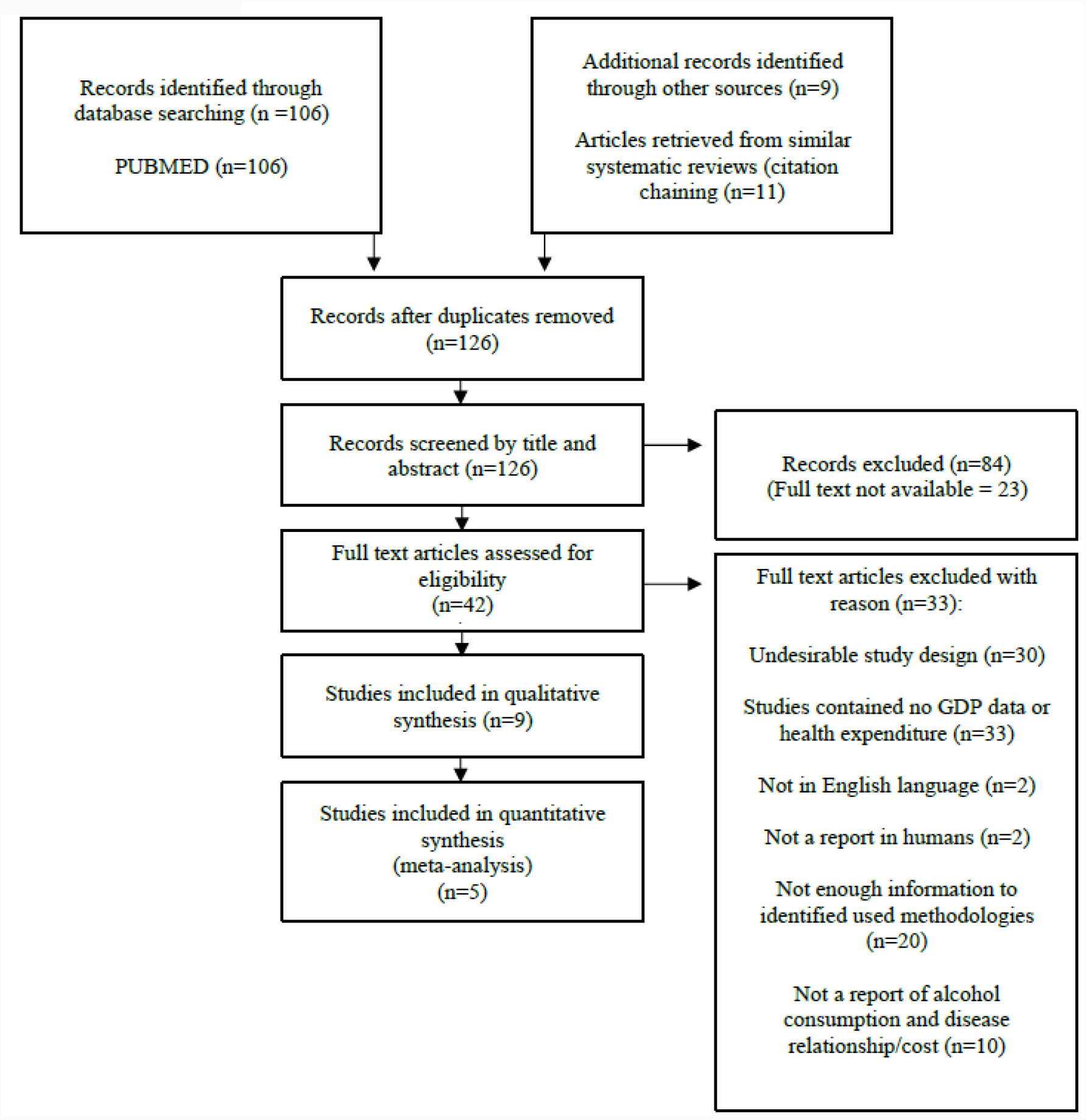
Prisma flowchart of studies included and excluded. A total of 9 studies were selected for the qualitative synthesis, however, only 5 studies were eligible for the meta-analysis due to the lack of sample information

### Characteristics of studies

Studies from 4 different regions were obtained: Asia (Thailand and Sri Lanka) (20,21), Africa (South Africa) (22), Europe (Germany, Sweden, Portugal, Belgium, and France) (23, 24, 25, 26, 27) and North America (Canada) (28) (Table 1). Although the studies observed have a varied geographical location, no studies from LICs were obtained. The proportion of HICs in this study is noticeably high, only one LMIC (Sri Lanka) was obtained. Of the 9 studies retrieved, 6 were of HICs (Germany, Sweden, France, Canada, Portugal, and Belgium), 2 studies were of UMIC (South Africa and Thailand) and the final study as aforementioned was of LMIC (Sri Lanka). All studies assessed the effect of alcohol consumption on health and healthcare costs. The Alcohol Attributable Fractions (AAF) played a key role in evaluating the economic effects of alcohol consumption on wellbeing. Data collection methods included surveys, self-reported drinking cases, the alcohol concentration in blood and administration records. This aided the estimations of alcohol-related health issues, deaths, accidents, road injuries, and crimes. The International Classification of Diseases (ICD-10/9) codes were used to express the disease class and associated figures (Table 2).

**Table 1:** Characteristics of selected studies. Year of estimates span from 1995-2015 across countries

| Author | Country | WDI classification | Data collection method | Sample size | Year of estimate costs | Study design | Gender | Cost (% of GDP) | Age | Alcohol related issues |
| --- | --- | --- | --- | --- | --- | --- | --- | --- | --- | --- |
| Elvira Lima and Teresa Esquerdo | Portugal | H | Secondary data | 19878 | 1995 | Cross-sectional | Male and female | 0.6 | >14 | Disease, injury and social |
| Fenoglio, Parel and Kopp | France | H | Secondary data | 58.7 million | 1997 | Cross-sectional | Male and female | 1.42 | n/a | Disease, injury and social |
| Jarl et al | Sweden | H | Survey | 3,000 | 2002 | Cross-sectional | Male and female | 1.13 | No limit | Disease, injury and social |
| Konnopka and König | Germany | H | Survey | 8139 | 2002 | Cross-sectional | Male and female | 1.16 | ≥18 | Disease, injury and social |
| Matzopoulos et al | South Africa | UM | Secondary data | n/a | 2009 | Cross-sectional | Male and female | 10-Dec | ≥18 | Disease, injury and social |
| Ranaweera et al | Sri Lanka | LM | Secondary data | n/a | 2015 | Cross-sectional | Male and female | 1.07 | n/a | Disease, injury and social |
| Rehm et al | Canada | H | Survey | n/a | 2002 | Cross-sectional | Male and female | 2.7 | No limit | Disease, injury and social |
| Thavorncharoensap et al | Thailand | UM | Survey | 3,029,427 | 2006 | Cross-sectional | Male and female | 1.99 | ≥15 | Disease, injury and social |
| Verhaeghe et al | Belgium | H | Survey and secondary data | Unclear | 2012 | Cross- sectional | Male and female | 0.4 |  | Disease, injury and social |

**Table 2:** Alcohol-related disease or where alcohol is a risk factor based on ICD-10 classification in studies countries.

| Author | Country | Disease class studied |
| --- | --- | --- |
| Elvira Lima and Teresa Esquerdo | Portugal | 2 3 4 5 6 7 8 9 10 |
| Fenoglio, Parel and Kopp | France | 2 3 4 5 6 7 8 9 10 |
| Jarl et al | Sweden | 2 3 4 5 6 7 8 9 10 |
| Konnopka and König | Germany | 2 3 4 5 6 7 8 9 10 |
| Matzopoulos et al | South Africa | 1 2 3 4 5 6 7 8 9 10 |
| Ranaweera et al | Sri Lanka | 2 3 4 5 6 7 8 9 10 |
| Rehm et al | Canada | 1 3 4 5 6 7 8 9 10 |
| Thavorncharoensap et al | Thailand | 1 2 3 4 5 6 7 8 9 10 |
| Verhaeghe et al | Belgium | 1 3 4 5 6 7 8 9 10 |

*Diseases have been categorized into 1= Infectious diseases, 2=Neoplasms, 3= Endocrine diseases,4=Psychiatric disorders, 5= Nervous system disorders, 6= Diseases of the circulatory system, 7= Digestive system diseases, 8= Skin conditions, 9= Pregnancy and childbirth related complications or issues within the prenatal period and 10= Injuries (accidental and non-accidental), poisoning*

The use of AAFs in these studies to estimate the fraction of diseases and consequent deaths which directly link to alcohol was imperative to ensure cost figures derived are specific to alcohol-related harm (24). The amount of alcohol consumed was expressed in grams per day. All studies included were observational cross-sectional study designs; all but one study used a prevalence-based approach in estimating the cost of illness (20-23, 25-28). From the studies analysed, it is the UMIC South Africa, that displayed the highest percentage of GDP, 10-12%, the question of whether infectious disease inclusion had an impact on this result is an outlet that needs to be explored. The study conducted by Rehm et al of Canada’s burden of alcohol presented its costs to equate to 2.7% of the GDP. Three of the countries which occupy the European region; Sweden, Germany and France, and shared similar values of alcohol-related costs, 1.42%, 1.16%, and 1.13% respectively. However, Belgium and Portugal displayed lower costs as a percentage of GDP, these being 0.4% and 0.6% correspondingly. Sri Lanka’s overall estimated value ranked lower than the majority (1.07%) whilst Thailand’s costs attributable to alcohol (1.99% of GDP) ranked third following South Africa and Canada.

All studies presented diseases that contributed to alcohol-related morbidity and mortality. Portugal’s biggest alcohol-related health concern was, as predicted, alcoholic cirrhosis. 82% of costs directly associated with alcohol use was attributable to the inpatient expenses of this disease. Alcoholic psychosis and alcohol dependence appeared to be the next alcohol-related health concern. In Sri Lanka, the total costs of the alcohol-related lip, oral and pharynx cancer (USD 42.16 million) and oesophagus cancer (USD 18.98 million) was considerably high. This in comparison to the total costs related to alcoholic liver cancer, which was USD 1.63 million is surprising, as liver cancer is the most known alcohol-related cancer in heavy drinkers. Approximately USD 95.63 million and USD 45.34 million was spent on alcoholic liver disease and alcohol use disorder respectively, these were the highest economic costs of the non-cancer disease group. The financial and medical costs of road injuries in Sri Lanka exceeds any alcohol-related disease; USD 251.28 million was put forward to cover the direct and indirect costs associated with this. A total of USD 28.28 million was associated with drowning; this was interesting to note; no studies in this analysis mentioned events such as this. Of particular relevance are the South African and Thai studies, the infectious diseases associated with alcohol are mentioned and included in the final cost estimations. The contribution of alcohol to infectious diseases included the human immunodeficiency viruses (HIV), tuberculosis and lower respiratory tract infections. These illnesses alongside violence and unintentional injury accounted for three-quarters of South Africa’s alcohol-related economic disease burden. Both tuberculosis and HIV/AIDS occupied the largest space of the DALY proportion, 18% and 13% respectively. Fenoglio, Parel and Kopp’s study on the French population stated 14% of males are addicted to alcohol, whilst this figure is nearly five times lower in the female population; only 3% are alcohol addicted (23). Fenoglio and colleagues not only studied the effects of alcohol but compared the associated statistics to tobacco and illicit drugs; it is evident that alcohol presents France with the greatest drug-related costs.

Table 3 displays the total net cost of alcohol misuse. In Thailand, 84.3% of the total costs were attributable to healthcare costs, this is closely followed by Germany where 83.7% are healthcare costs. Sweden’s healthcare costs like many studies in this paper included in-patient and out-patient costs, medical care, co-morbidity, pharmaceuticals, and some studies also included employee assistance programs (EAPs) which were covered by employers. Thailand expressed the greatest healthcare cost percentage out of all observed studies; the evidence surrounding the relationship between alcohol and infectious diseases has been expanding in recent times. Hence Thavorncharoensap et al’s study of Thailand included the costs associated with unsafe sexual practises and therefore estimates included HIV/AIDS cases, this, therefore, contributes to the higher percentage in healthcare spending. The healthcare costs in Canada totalled USD $3,306.2 million, a heavy fraction of this is traceable to acute care hospitalisation, inpatient specialised treatment, and prescription drugs. The costs associated with psychiatric hospitalisation was considerably low, in comparison to findings of that in Sri Lanka; this is noteworthy.

**Table 3:**
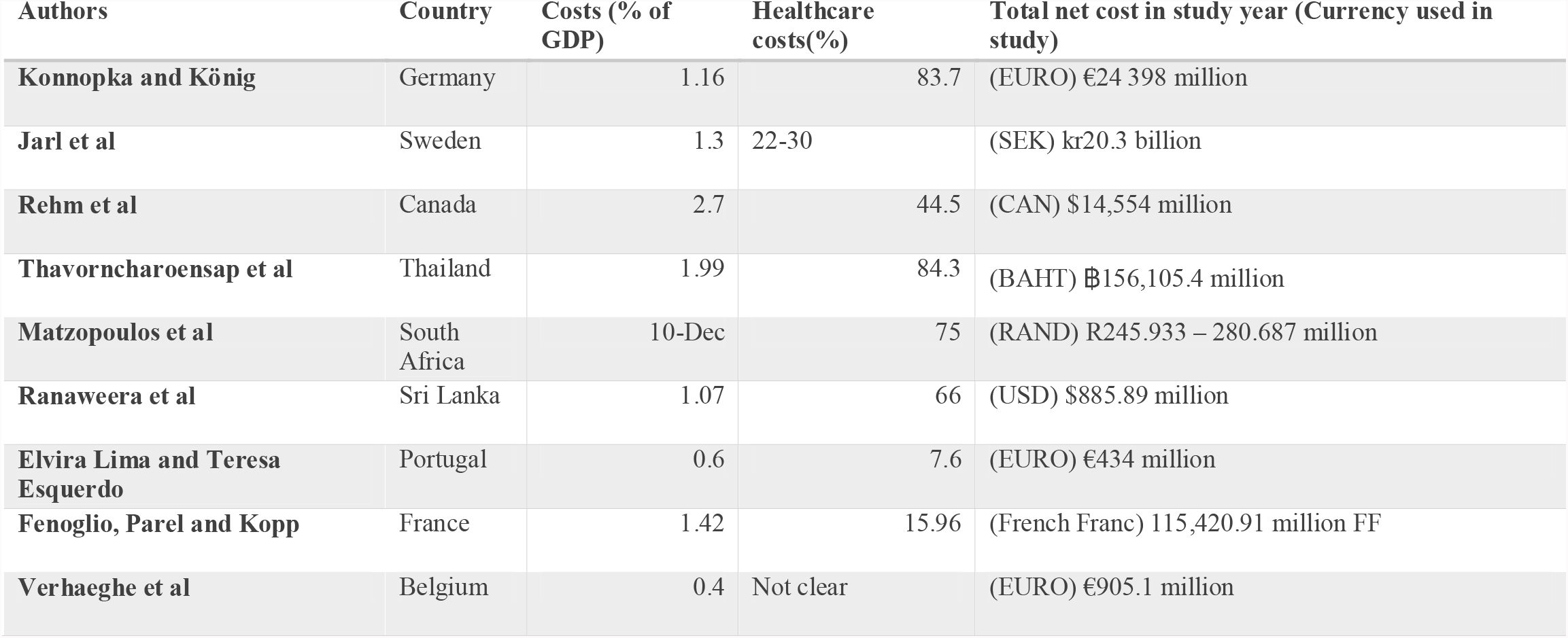
The estimated percentage of GDP attributable to alcohol costs, total costs and proportion of costs spent on alcohol related care treatment

The percentage of costs attributable to direct and indirect costs is demonstrated in Table 4. The specific estimates for the direct and indirect costs of the South African study were not clear and thus were not tabulated. Germany, Sweden, Thailand, Sri Lanka, and Portugal all shared the same results with regards to having a higher percentage of indirect costs than direct costs, however, this was the opposite for countries such as Belgium, France, and Canada. As seen before, Thailand’s greatest direct cost is healthcare costs, followed by law enforcement costs such as court and police costs. Canada’s largest direct cost was direct law enforcement costs, this was over two times the cost of acute hospitalisation. Unlike the figures seen in Canada regarding social costs, the direct costs implementing the law in France were some of the lowest expenses. 83% of Portugal’s direct costs were made up of acute general hospitals. Financial costs associated with psychiatric hospitals only accounted for 1% in Portugal. In stark contrast lies Belgium’s largest direct cost, psychiatric hospitals; two times the cost put forward to cover general hospital costs. The largest indirect costs in Sweden were for lost productivity due to mortality and long-term sickness, the same outcomes were seen in Germany’s indirect costs. It was interesting to note that in France, the greatest indirect cost was company losses in production and losses of income for private agencies. Premature mortality was a leading indirect cost in Sri Lanka (USD 388.86 million); the total cost of premature mortality equated to the complete total direct cost (USD 388.39 million).

**Table 4:** Percentage of direct and indirect costs attributable to negative consequences of alcohol consumption.

| Author | Country | Direct costs % | Indirect costs (%) |
| --- | --- | --- | --- |
| Elvira Lima and Teresa Esquerdo | Portugal | 25.1 | 74.9 |
| Fenoglio, Parel and Kopp | France | 50.2 | 49.8 |
| Jarl et al | Sweden | 37 | 63 |
| Konnopka and König | Germany | 35 | 65 |
| Matzopoulos et al | South Africa | - | - |
| Ranaweera et al | Sri Lanka | 44 | 66 |
| Rehm et al | Canada | 51 | 49 |
| Thavorncharoensap et al | Thailand | 4 | 66 |
| Verhaeghe et al | Belgium | 58.5 | 41 |

A total of 5 studies were used in the meta-analysis. The remaining 4 studies could not be computed due to the lack of sample size information. The results of the random effects meta-analysis included studies conducted in Germany, Sweden, Portugal, France, and Thailand; 4 from the European region and one from Asia (Figure 3 and 4). The restricted maximum likelihood estimator calculated the tau^2 to be 0 (SE=0.0002) and heterogeneity was estimated to be 0.00%. Outcomes reflected the notion that there was no variation which reflected differences in the mean population. The confidence interval test revealed a 95% confidence interval (0.00 and 98.56). The test for heterogeneity revealed a p-value of 0.447, which hints at insignificant heterogeneity. The I^2 value suggests homogeneity across all studies and hence no moderator analysis and subsequent baujat plot were required in this instance. A pooled estimate model coefficient (summary effect size with standard error) was shown to be 0.01%.

**Figure 3:**
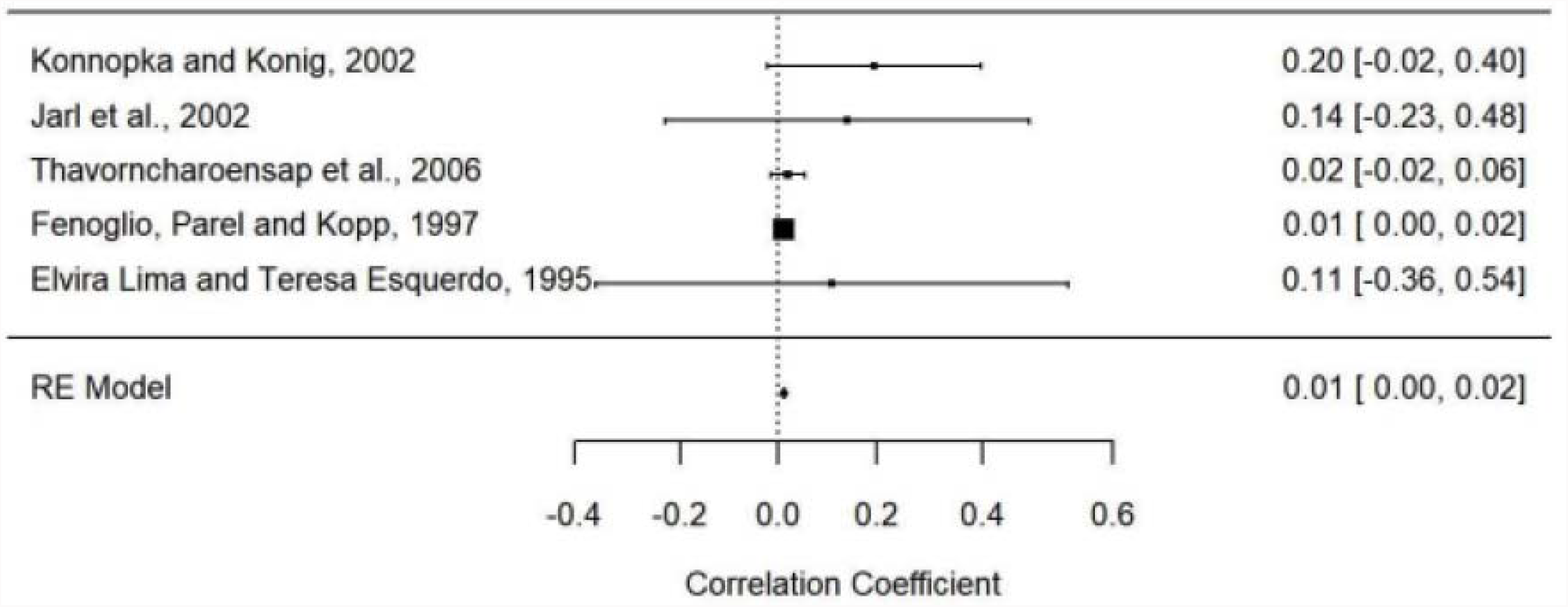
**Results of the forest plot**, homogeneity detected amongst all studies. Results suggest alcohol negatively affects society though in this period, not substantially.

**Figure 4:**
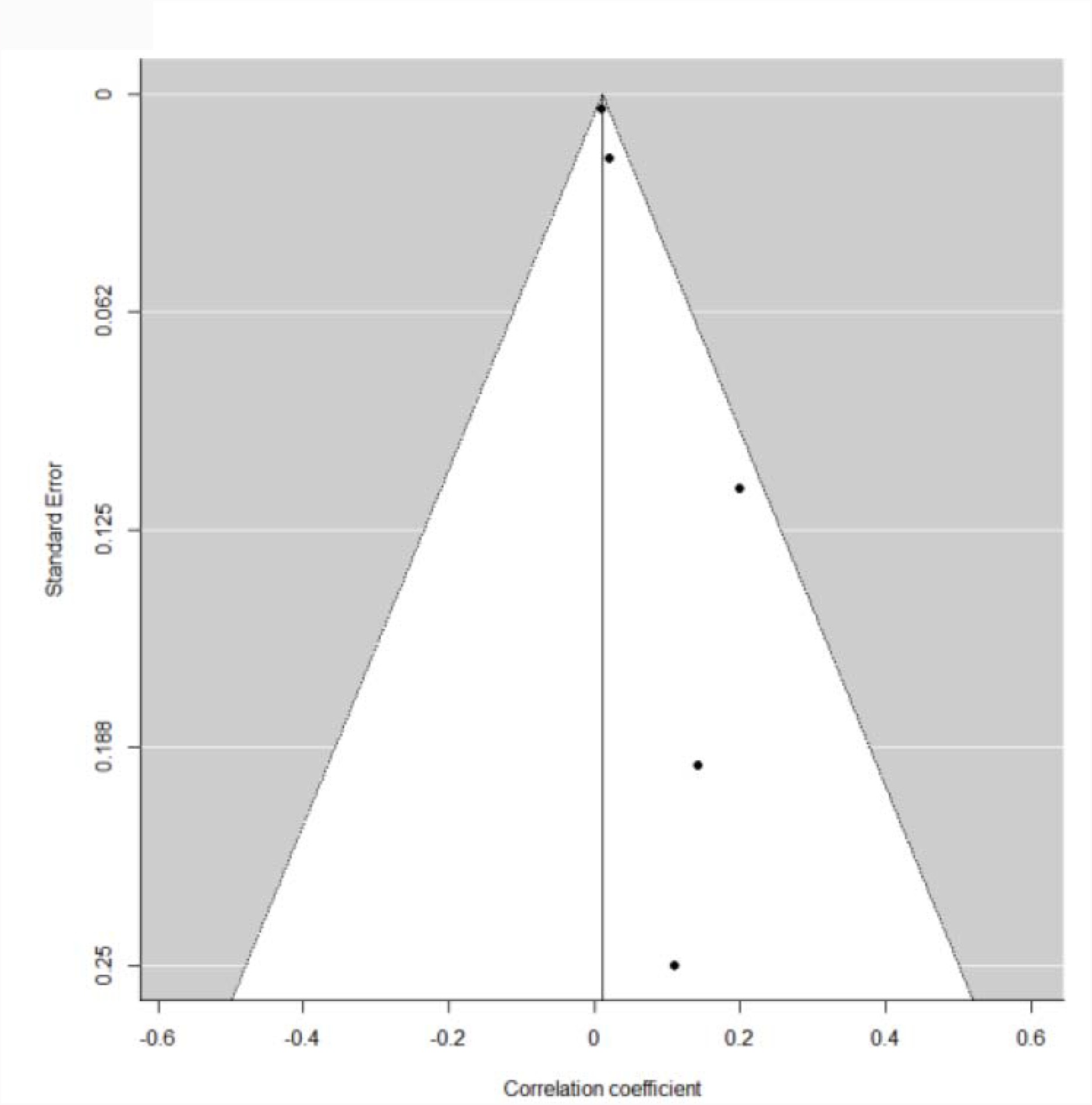
**Funnel plot** showing asymmetry in study, contradicted by the results of the homogeneity test. A possible explanation for this is the lack of negative results in studies as all outcomes were measured as a percentage. The rank correlation test and Egger’s regression test was conducted to confirm the accuracy of the funnel plot, no bias detected.

## DISCUSSION

We found that albeit the pooled estimate for the economic burden of alcohol consumption of the selected 5 studies was small (0.01%); it is nevertheless suggestive that alcohol is destructive to both physical and economic health. The findings of this study revealed that generally, most alcohol-related expenditure was put towards covering acute hospitalisation and law enforcement costs. Although there are two systematic reviews available assessing the economic burden of alcohol, this current report is unique; the meta-analysis carried out in this project provides a more precise estimate of the effect. One of the systematic reviews, conducted by Thavorncharoensap et al, estimated that the economic burden of alcohol consumption in 12 countries, of which 11 were HICs and one a UMIC, was 0.45%-5.44% (15). Whilst Baumberg’s investigation evaluated the global GDP attributed to alcohol costs was 0.6-2.0%; however, this is hugely inaccurate due to all 16 studies being on HICs (7).

The major alcohol-related disease across the observed countries were AUDs, liver disease, alcohol-related cancer, and road injuries. From the data provided, we can infer that increasing awareness surrounding alcohol addiction and perhaps increasing efforts to persuade individuals who need mental health attention to seek help could potentially alleviate the AUD burden and thus reduce the number of disease cases associated with chronic and binge drinking (29). Men generally drink more alcohol, this social behaviour is seen as making one seem more masculine, whilst women, especially in developing countries tend to shy away from drinking, possibly due to their stronger commitment to religious beliefs and community expectations (30). This gender difference is seen in the French study, the proportion of males addicted to alcohol was almost five times more than females. Improving mental health application amongst the male population could reduce national health costs; we can expect to see a decrease in disease cases where alcohol is a direct or component cause, and as a result anticipate a decline in treatment costs and costs to society (30, 31).

In France and other HICs, the prevalence of alcohol consumption is elevated. This is observed in Fenoglio, Parel and Kopp’s study which highlighted alcohol as being the country’s major drug problem. Of course, with France possessing one of the highest nominal GDPs across the globe, civilians can afford this psychoactive drug, enjoy the short-lived luxuries of it, and may suffer the long-term consequences (32). However, in places like South Africa, a large proportion of its residents live in poverty. The relationship between lower socioeconomic statuses (SES) and ethanol consumption is noteworthy; low-income earners in UMIC, LMIC and LICs adopt more dangerous drinking patterns (33). In such a setting, combined with the vast inequalities in health systems and public schemes seen in places like South Africa and Sri Lanka, lay a foundation for calamity. With a lack of education being provided to locals, the alcohol industry is seeing success through their marketing approaches, especially with the younger population and women.

Research into the relationship between alcohol consumption and infectious disease is new; with most of the global population living in developing countries and a high proportion of the global burden of disease being preventable infectious diseases, this area of study calls for rapid growth and financial resources (34). Our study included two papers in which the author included infectious disease costs in their final cost estimations, these studies were conducted in Thailand and South Africa. Surprisingly, these countries displayed the highest healthcare costs associated with alcohol, with South Africa showing the greatest percentage of GDP spent on alcohol-related costs. It can be thus suggested that although it is known that the prevalence of sexually transmitted diseases (STDs) is much higher in LIC and LMIC countries due to the lack of education, it is further accentuated by alcohol intake (35). Early teaching about the major contributors to health inequalities, mental health and STDs should be an utmost priority in the agendas of many countries, especially those of low-income (36). Owing to globalisation, and as transnational companies continue to shift their focus onto developing countries, low-income nations can expect to see an upsurge in the number of alcohol consumers and thus medical and social costs; overlooking these concerns will invite not only poor health but an even poorer nation (37).

The WHO has recognised that alcohol consumption may become a greater problem in years to come, one of their responses are the “best buys”. This consists of promoting bans on alcohol advertisement, limiting alcohol availability, and increasing alcohol taxes (38). Even so, this may not suffice. The International Agency of Research on Cancer has identified alcohol as a carcinogen (39). As a consequence of inadequate funding, disappointments in political motivation, and policymaking hindrances from the alcohol industry; developing countries cannot make progress in implementing effective policies without the aid of national and international organisations (37). An overt national policy is needed in some LICs with some HICs in need of regional schemes regarding alcohol consumption (38,37). The ease of access to quality and affordable rehabilitation and treatment centres in LICs needs to be considered to address the high proportion of mental illnesses in developing countries. In aid to upscale alcohol policies, the launch of a WHO cabinet such as the Tobacco-free initiative may be a promising retort (37).

Our study, as well as the two other systematic reviews conducted in this field, did not have access to alcohol-related figures in LICs and many LMICs. The lack of research in these countries is worrying, and unless developing countries train individuals to become producers of research, the progress of the country’s health will be impeded. The measurement of externalities is a gap in research that needs attention (40). Some studies included in this paper used self-reported data for alcohol consumption statistics; this introduces uncertainty about the level of accuracy in final cost estimations due to these types of data collection methods having miscalculations regarding the proportion of alcohol consumed by individuals (19, 26). Therefore, the pooled estimate derived from this study is more than likely an underestimate; the economic harm present in developing countries is requisite for proper calculations.

In LICs data with geotagged information concerning suppliers and policy information may not be recorded (33). In these settings, the difficulty that accompanies accessing government information, as well as the poor research capacity in these countries, need to be overcome. Researching LICs could provide invaluable knowledge to offer solutions to lower the high premature mortality rates seen across the world (34). However, for research to be conducted in such places, foreign support is required. All studies in the report used a prevalence-based approach in their methods, and though this is useful when understanding needs for government spending, this may not be informative when measuring the impact of alcohol policy. However, there is again a paucity in available studies that use an incidence-based approach in understanding the effects of alcohol policy (20). To fully alert leaders in public health as well as governments, NGOs and IGOs, more studies conducted in HICs and LICs using a human capital approach is an obligation to present an estimate for future and present costs related to alcohol mortality (41).

This study demonstrated that alcohol use, more precisely the unsafe practice of alcohol use, is becoming a major public health concern in developed and developing countries. Not only is there a plethora of evidence to illustrate the link between ethanol consumption and non-communicable diseases, but there is a growing body of research with regards to alcohol being a risk factor for the contraction of infectious disease (42). The available literature surrounding alcohol consumption in HICs is expanding in recent times, though studies on the effects of alcohol in LICs is extremely sparse (43). With the alcohol marketing industry encouraging the youth to adopt harmful lifestyle practices, countries still undergoing economic development have become targets to aid the growth of the alcohol consumer base (36). Researchers preparing to conduct future research in this field should consider finding solutions to minimize the possibility of encountering the limitations discussed in this study. Furthermore, future work involving cost of illness studies should aim to assess the effect of socioeconomic variables, such as education and income, on cost results by incorporating subgroup analyses in their research investigations. Additionally, future studies also should consider carrying out research projects that aim to assess alcohol costs that are preventable and use a human capital approach. Adequate funding is needed in order to access high quality accurate data; appropriate resource allocation will not only provide invaluable scientific evidence regarding alcohol consumption but allow more effective policy making and interventions (34).

This study has provided an estimate for the proportion of GDP attributable towards the negative effects of alcohol consumption. Crucial information required by public health professionals and researchers in aiding policies, infrastructure and interventions has been revealed in the current paper. This report sternly recommends prioritising the obtainment of financial aid required for research in low-income countries to undoubtedly provide a more precise estimate for the global GDP ascribable to alcohol consumption.

## Data Availability

All data used for the analysis of this systematic review is presented within the manuscript.

## ACKNOWLEGDMENTS

R.P. holds a fellowship supported by Rutherford Fund from Medical Research Council (MR/R0265051/1 & MR/R0265051/2).

## Patient consent for publication

Not required.

## Ethical approval

This is a systematic review and meta-analysis and therefore involves published data. Respective researchers did not have access to individual patient information, thus the potential for ethical issue was not raised in this research. Therefore, the requirement for ethical approval and patient consent was waived.

